# Neighborhood Socioeconomic Status and Short-Term Postoperative Complications Following Bariatric Surgery

**DOI:** 10.1101/2025.09.16.25335899

**Authors:** Oluwasegun Akinyemi, Terrence Fullum, Mojisola Fasokun, Oluebubechukwu Eze, Kakra Hughes, Dahai Yue, Craig Scott Fryer, Jie Chen, Kelle White Whilby

**Author notes:** Corresponding author: Oluwasegun Akinyemi.

## Abstract

**Background:** Obesity is a significant public health concern, affecting over 41.9% of adults in the United States and contributing to increased risk of chronic conditions and premature mortality. Bariatric surgery is the most effective long-term intervention for severe obesity, offering sustained weight loss and improved metabolic outcomes. However, socioeconomic disparities persist in both the utilization and postoperative outcomes of bariatric surgery, particularly among those who live in socioeconomically disadvantaged neighborhoods.

**Objective:** This study evaluates the impact of neighborhood socioeconomic status (nSES), as measured by the Distressed Communities Index (DCI), on short-term postoperative complications following bariatric surgery. Additionally, we investigate whether race/ethnicity moderates this association.

**Methods:** We conducted a retrospective cohort study using data from the Maryland State Inpatient Database (SID) from 2018 to 2020. The study population included adult patients (≥18 years) who underwent bariatric surgery during the study period. The primary outcome was the occurrence of short-term postoperative complications (e.g. gastrointestinal leaks, gastrointestinal bleeding and venous thromboembolism) categorized using a composite complication variable defined as no complications or one or more complication. The primary independent variable was nSES, classified into five categories based on the DCI (prosperous, comfortable, mid-tier, at-risk, and distressed). Multivariable ordinal logistic regression models were used to assess the association between nSES and the occurrence of one or more complication post-surgery, adjusting for demographic, clinical, and socioeconomic and geographic covariates. We tested whether race/ethnicity modified the association between nSES and the selected outcomes.

**Results:** Among 10,784 patients who underwent bariatric surgery in the study period, 94.7% had no postoperative complications, 5.3% experienced one or more complications. The most common complications were venous thromboembolism (2.2%) and gastrointestinal leaks (0.9%). Multivariable analyses revealed no statistically significant association between nSES and the occurrence of short-term postoperative complications. Additionally, race/ethnicity did not significantly moderate this relationship.

**Conclusions:** This study found no significant association between neighborhood socioeconomic disadvantages and the occurrence of short-term postoperative complications following bariatric surgery.

## INTRODUCTION

Obesity is a significant public health challenge in the United States, affecting approximately 41.9% of adults and contributing to increased risk of chronic conditions such as diabetes, cardiovascular disease, and certain cancers and premature mortality [1–3]. The economic burden of obesity continues to rise, with estimates exceeding $347.5 billion annually [4]. Bariatric surgery has emerged as the most effective long-term intervention for severe obesity (BMI ≥ 40 kg/m² or BMI ≥ 35 kg/m² with comorbidities), offering sustained weight loss, improved metabolic outcomes, and reduced mortality [5, 6]. Over the past two decades, bariatric surgery has evolved with improved techniques and safety profiles, leading to a steady increase in its utilization [7–9]. In 2017 alone, approximately 228,000 bariatric procedures were performed in the United States [8]. Advances in surgical protocols, anesthesia, and post-operative care have significantly reduced complication rates, making bariatric surgery safer and more accessible [10]. However, despite these advancements, socioeconomic disparities in utilization and outcomes persist.

Although bariatric surgery offers significant benefits, short-term post-operative complications remain a concern [11, 12]. Common complications include gastrointestinal (GI) leaks, surgical site infections (SSIs), and venous thromboembolism (VTE), all of which can lead to increased morbidity, prolonged hospital stays, readmissions, and reduced patient satisfaction [13, 14]. These complications may discourage eligible patients from pursuing surgery due to concerns about post-operative recovery and financial costs [15]. Patients from disadvantaged communities may be particularly at risk for these adverse outcomes due to systemic barriers to high-quality post-operative care, a higher prevalence of pre-existing comorbid conditions, and other social determinants of health that may impact post-surgical recovery [16, 17]. Understanding the factors that contribute to these disparities is essential to improving surgical outcomes and increasing equitable access to bariatric surgery.

Several studies have underscored the clinical and public health implications of post-operative complications following bariatric surgery [11, 18–20]. For instance, gastrointestinal leaks, though relatively rare, can occur in up to 5% of patients and are associated with significant morbidity and mortality [21]. Surgical site infections affect approximately 0.4% to 12% of patients, depending on the procedure type and hospital protocols [22]. Venous thromboembolism (VTE) remains a leading cause of post-operative morbidity despite prophylaxis, with reported incidence rates ranging from 0.2% to 1.3% [23]. Importantly, the risk of these complications may be amplified in patients from low socioeconomic status (SES) neighborhoods, where limited healthcare resources, lower health literacy, reduced social support, and under-resourced post-acute care settings can hinder timely recognition and management of early signs of complications [17, 24]. These neighborhood-level structural barriers can compound individual vulnerabilities, increasing the likelihood of adverse outcomes and potentially reinforcing long-standing disparities in surgical recovery and overall health status [24, 25].

Social determinants of health (SDOH), including individual-level socioeconomic status (SES), race, insurance type, and income, play a critical role in post-surgical outcomes [26]. Studies have shown that individuals with lower SES or public insurance experience higher rates of complications, prolonged hospital stays, and readmissions following bariatric surgery [26]. These disparities are often attributed to variations in access to high-quality healthcare, lower rates of post-operative follow-up, and differential exposure to environmental and lifestyle risk factors [17, 27]. While much research has focused on individual-level socioeconomic factors, there is growing recognition that neighborhood socioeconomic status (nSES) may offer additional predictive value in understanding socioeconomic disparities in surgical outcomes [28, 29].

Neighborhood socioeconomic status captures broader structural and environmental factors that influence health beyond individual-level SES [30–32]. Lower nSES neighborhoods are characterized by higher poverty rates, lower education levels, reduced access to healthcare services, food insecurity, and higher levels of chronic stress [33, 34]. These factors contribute to disparities in post-operative complications and hospital readmissions among bariatric surgery patients [17, 34]. Research suggests that individuals from socioeconomically distressed neighborhoods face greater barriers to healthcare access, which may translate to delayed diagnosis and treatment of complications, poorer adherence to post-operative care, and worse overall outcomes [17, 33]. However, there is limited research on how nSES, as measured by indices such as the Distressed Communities Index (DCI), influences short-term post-operative complications following bariatric surgery.

Race and ethnicity are established factors that influence health outcomes, and their interaction with neighborhood socioeconomic status may further exacerbate the observed disparities [35, 36]. Black and Hispanic patients undergoing bariatric surgery have been found to experience higher rates of complications compared to White patients, even after adjusting for individual SES [16, 36]. Structural inequities in healthcare delivery, differential referral patterns, and provider biases contribute to these disparities [36, 37]. Given that racial and ethnic minority populations are more likely to reside in low-nSES neighborhoods, it is crucial to investigate whether race/ethnicity moderates the association between nSES and short-term post-operative complications [37, 38]. This study aims to address this gap by examining whether nSES, as measured by the DCI, is associated with short-term post-operative complications after bariatric surgery, including gastrointestinal leaks, surgical site infections, and venous thromboembolism (VTE). This study will also examine whether race/ethnicity moderates this association. Understanding these relationships is essential to improving healthcare equity and optimizing bariatric surgery outcomes for patients from disadvantaged communities.

## METHODOLOGY

### Study Design and Population

This study employs a retrospective cohort design using data from the Maryland State Inpatient Database (SID) from 2018 to 2020 [39]. The study population includes all adult patients (≥18 years) who underwent bariatric surgery in Maryland during the study period. Bariatric procedures were identified using ICD-10 procedure codes for sleeve gastrectomy, gastric bypass, adjustable gastric banding, and other recognized bariatric surgeries. The Maryland SID provides a comprehensive, statewide dataset that captures hospital discharges across all payers, ensuring reduced bias related to insurance status.

### Primary Outcome: Short-Term Post-Operative Complications

The primary outcome of interest was the occurrence of one or more short-term post-operative complications following bariatric surgery. This outcome was defined as a binary variable: individuals who did not experience any short-term complication were coded as 0, while those who experienced at least one complication were coded as 1. To construct this variable, we included the most commonly reported short-term post-operative complications in the literature.

The selected complications included gastrointestinal leaks, stenosis, gastrointestinal bleeding, venous thromboembolism (VTE), gastroesophageal reflux disease (GERD), pouch stress, persistent vomiting, post-operative pain, acute cholecystitis, and in-hospital mortality. These complications were chosen based on their documented prevalence and clinical relevance as early adverse outcomes among bariatric surgery patients [40–43].

In addition to this binary composite measure, we also examined the association between neighborhood socioeconomic status (as measured by the Distressed Communities Index, DCI) and individual short-term complications, specifically, gastrointestinal leaks, venous thromboembolism (VTE), and gastrointestinal leaks.

### Main Explanatory Variable: Neighborhood Socioeconomic Status (nSES)

The main independent variable is nSES, captured using the DCI quintiles. The DCI is a ZIP-code-based measure developed by the Economic Innovation Group, integrating seven economic indicators (e.g., unemployment rates, poverty levels, housing vacancy rates, and business vitality) to assess socioeconomic distress. For this study, the DCI is categorized into five quintiles: Quintile 1 (Prosperous), Quintile 2 (Comfortable), Quintile 3 (Mid-tier), Quintile 4 (At-risk), Quintile 5 (Distressed).

Higher quintiles represent greater socioeconomic disadvantages, allowing for a dose-response analysis of nSES and postoperative complications.

### Covariates

To account for potential confounding, the study will adjust for key demographic, clinical, socioeconomic and geographic factors, including:

Demographic Variables: Age (continuous), sex (male/female), race/ethnicity (White, Black, Hispanic, and Other).

Clinical Variables: Charlson commodity index (CCI) is a weighted scoring system that quantifies the burden of comorbid conditions based on their severity and association with mortality, with higher scores indicating greater comorbidity burden and risk of adverse health outcomes) and obesity severity (BMI ≥35 kg/m² with comorbidities or BMI ≥40 kg/m²) [44–48].

Socioeconomic Factors: Insurance type (Medicare, Medicaid, Private, Self-pay), and household median income (categorized into quintiles based on census data).

### Geographical Factors

The Urban/Suburban/Small Town/Rural designation variable (Urban) categorizes patients based on their residential area type. This numeric categorical variable ranges from 1 to 4, representing the following classifications: Rural (1), Small Town (2), Suburban (3), and Urban (4). This variable was included in the analysis to examine potential geographic disparities in bariatric surgery utilization based on urbanicity and rurality.

### Statistical Analysis

Descriptive analyses were conducted to summarize baseline characteristics by the occurrence of short-term post-operative complications or otherwise. Categorical variables were compared using χ² tests, while continuous variables were assessed using either t-tests or Mann-Whitney U tests, as appropriate.

To evaluate the association between nSES and the occurrence of short-term post-operative complications, multivariable logistic regression models were employed. The binary outcome variable indicated whether a patient experienced any short-term complication (1) or none (0) following bariatric surgery. The primary explanatory variable was the DCI, used as a measure of nSES. Covariates were included in a stepwise fashion across models to assess the robustness of the association. The logistic regression model was specified as follows:

Logit(P(Y=1))=α+β1(DCI)+β2(Covariates)

Given well-documented racial and ethnic disparities in surgical outcomes, race/ethnicity was tested as a potential effect modifier in the association between DCI and post-operative complications. An interaction term between DCI and race/ethnicity was included in a separate model to assess whether the relationship varied across racial groups:

Logit(P(Y=1))=α+β1(DCI)+β2(Covariates)+β3(Race)+β13(Race×DCI)

The interaction term (β₁₃) captured whether the effect of DCI on the likelihood of experiencing a post-operative complication differed by race/ethnicity. A statistically significant interaction (p < 0.05) was interpreted as evidence of effect modification.

### Sensitivity Analyses

To ensure the robustness of the results, sensitivity analyses were conducted using alternative approaches to measure neighborhood socioeconomic status (nSES) and address potential confounding. The Area Deprivation Index (ADI) was used as an alternative measure of nSES to assess whether the findings were consistent across different socioeconomic indices.

### Ethical Considerations

This study utilizes de-identified secondary data from the Maryland SID, exempt from direct patient consent under HIPAA regulations. The study protocol was reviewed and approved by the University of Maryland Institutional Review Board (2284677-1).

## RESULT

### Baseline Characteristics

Table 1 presents the demographic, socioeconomic, and clinical characteristics of the study population, stratified by the occurrence of any short-term post-operative complication following bariatric surgery. Among the 10,784 individuals included in the analysis, 566 (5.3%) experienced at least one short-term complication, while 10,218 (94.8%) did not.

**Table 1.** Baseline Characteristics of Bariatric Surgery Patients in Maryland by Postoperative Complication Status, 2018–2020.

| **Variable** | **Total Population**<br>**(N=10,784)** | **Complication**<br>**(n=566)** | **None (n=10,218)** | **Chi**<br>**Squared** | **p-**<br>**value** |
| --- | --- | --- | --- | --- | --- |
| **Distressed**<br>**Communities Index** |  |  |  | 2.395 | 0.664 |
| Prosperous | 2,645 (24.5%) | 149 (26.3%) | 2,496 (24.4%) |  |  |
| Comfortable | 3,085 (28.6%) | 164 (29.0%) | 2,921 (28.6%) |  |  |
| Mid-Tier | 2,204 (20.4%) | 117 (20.7%) | 2,087 (20.4%) |  |  |
| At-Risk | 1,487 (13.8%) | 74 (13.1%) | 1,413 (13.8%) |  |  |
| Distressed | 1,363 (12.6%) | 62 (11.0%) | 1,301 (12.7%) |  |  |
| **Age (mean + std)** | 44.1 + 11.6 | 49.1 + 12.3 | 43.8 + 11.5 | t=-10.650 | <0.001 |
| **Age (Yr.)** |  |  |  | 119.224 | <0.001 |
| 18-44Yr. | 5,656 (52.5%) | 206 (36.4%) | 5,450 (53.3%) |  |  |
| 45/64Yr. | 4,638 (43.0%) | 290 (51.2%) | 4,348 (42.6%) |  |  |
| >64Yr. | 490 (4.5%) | 70 (12.4%) | 420 (4.1%) |  |  |
| **Female (ref. Man)** | 8,972 (83.2%) | 452 (79.9%) | 8,520 (83.4%) | 4.763 | 0.029 |
| **Race/Ethnicity** |  |  |  | 5.096 | 0.165 |
| White | 4,834 (44.8%) | 278 (49.1%) | 4,556 (44.6%) |  |  |
| Black | 5,209 (48.3%) | 253 (44.7%) | 4,956 (48.5%) |  |  |
| Hispanic | 503 (4.7%) | 26 (4.6%) | 477 (4.7%) |  |  |
| Other | 238 (2.2%) | 9 (1.6%) | 229 (2.2%) |  |  |
| **Insurance** |  |  |  | 113.751 | <0.001 |
| Self-Pay | 33 (0.3%) | 4 (0.7%) | 29 (0.3%) |  |  |
| Medicare | 975 (9.0%) | 120 (21.2%) | 855 (8.4%) |  |  |
| Medicaid | 2,087 (19.4%) | 80 (14.1%) | 2,007 (19.7%) |  |  |
| Private | 7,549 (70.0%) | 356 (62.9%) | 7,193 (70.4%) |  |  |
| Other | 138 (1.3%) | 6 (1.1%) | 132 (1.3%) |  |  |
| **CCI** |  |  |  | 41.660 | <0.001 |
| CCI =0 | 3,240 (30.0%) | 120 (21.2%) | 3,120 (30.5%) |  |  |
| CCI= 1-2 | 3,631 (33.7%) | 172 (30.3%) | 3,459 (33.9%) |  |  |
| CCI >2 | 3,913 (36.3%) | 274 (48.4%) | 3,639 (35.6%) |  |  |
| **Obesity**<br>**Classification** |  |  |  | 4.822 | 0.028 |
| Class I Obesity | 2,344 (21.7%) | 144 (25.4%) | 2,200 (21.5%) |  |  |
| Class II Obesity | 8,440 (78.3%) | 422 (74.6%) | 8,018 (78.5%) |  |  |
| **Urban** |  |  |  | 6.064 | 0.109 |
| Rural | 971 (9.0%) | 65 (11.5%) | 906 (8.9%) |  |  |
| Small Town | 744 (6.9%) | 44 (7.8%) | 700 (6.9%) |  |  |
| **Suburban** | **7,281 (67.5%)** | **373 (65.9%)** | **6,908 (67.6%)** |  |  |
| Urban | 1,788 (16.6%) | 84 (14.8%) | 1,704 (16.7%) |  |  |

The mean age of the overall cohort was 44.1 ± 11.6 years. Patients who experienced complications were significantly older (49.1 ± 12.3 years) compared to those without complications (43.8 ± 11.5 years; p < 0.001). Age group distribution also differed significantly (p < 0.001), with the highest complication rate among patients aged 45–64 years (51.2%), followed by those 18–44 years (36.4%) and those ≥65 years (12.4%).

Women comprised 83.2% of the total sample, with a slightly lower representation among those with complications (79.9%) compared to those without (83.4%; p = 0.029). While the racial and ethnic distribution did not reach statistical significance (p = 0.165), White patients had a slightly higher proportion in the complication group (49.1%) compared to those without complications (44.6%). Black patients made up 48.3% of the total cohort, followed by Hispanic (4.7%) and Other race (2.2%).

Insurance type was significantly associated with complication status (p < 0.001). Patients with Medicare were more likely to experience complications (21.2%) than those without (8.4%), while those with private insurance were less likely to experience complications (62.9%) compared to their counterparts without complications (70.4%).

Comorbidity burden, as measured by the Charlson Comorbidity Index (CCI), was significantly associated with complications (p < 0.001). Nearly half of the patients with complications (48.4%) had a CCI >2, compared to 35.6% among those without complications. Patients with no comorbidities (CCI = 0) were less likely to experience complications (21.2% vs. 30.5%).

Obesity classification was also associated with complication risk (p = 0.028). Class I obesity was more common among patients with complications (25.4%) compared to those without (21.5%). Class II obesity was slightly less prevalent among those with complications (74.6%) compared to the non-complication group (78.5%).

There were no statistically significant differences in complication status by neighborhood socioeconomic status (DCI quintiles; p = 0.664) or urban-rural residence (p = 0.109), though minor variations were observed across geographic categories.

### Incidence of Short-term Postoperative Complications

Table 2 presents the incidence of short-term postoperative complications among patients who underwent bariatric surgery. Among the 10,784 individuals who underwent bariatric surgery, 566 (5.3%) experienced at least one short-term postoperative complication (Table 2). The most common complication was venous thromboembolism, occurring in 2.2% of patients.

**Table 2:** Short-Term Complication Rates Post-Bariatric Surgery.

| **Complications (n=566)** | **Frequency** | **Percentages (%)** |
| --- | --- | --- |
| All Complications | 566 | 5.30% |
| **Commonest Complications** |  |  |
| Venous Thromboembolism | 238 | 2.20% |
| Gastrointestinal Leak | 94 | 0.90% |
| Gastrointestinal Bleeding | 62 | 0.60% |

Gastrointestinal leak and gastrointestinal bleeding were also notable, with rates of 0.9% and 0.6%, respectively. These findings highlight the relatively low but clinically significant risk of adverse events following bariatric surgery.

### Adjusted Odds of Postoperative Complications

After adjusting for relevant covariates, there were no statistically significant differences in the odds of experiencing any postoperative complication across DCI quintiles when compared to patients from Prosperous communities (Table3). Patients from Distressed communities had a lower, though non-significant, odds of developing any complication (aOR: 0.81; 95% CI: 0.56– 1.17), while those from Comfortable (aOR: 0.96; 95% CI: 0.76–1.21), Mid-Tier (aOR: 1.01; 95% CI: 0.78–1.30), and At-Risk (aOR: 0.93; 95% CI: 0.69–1.25) communities showed similar non-significant associations (Table 3).

**Table 3:**
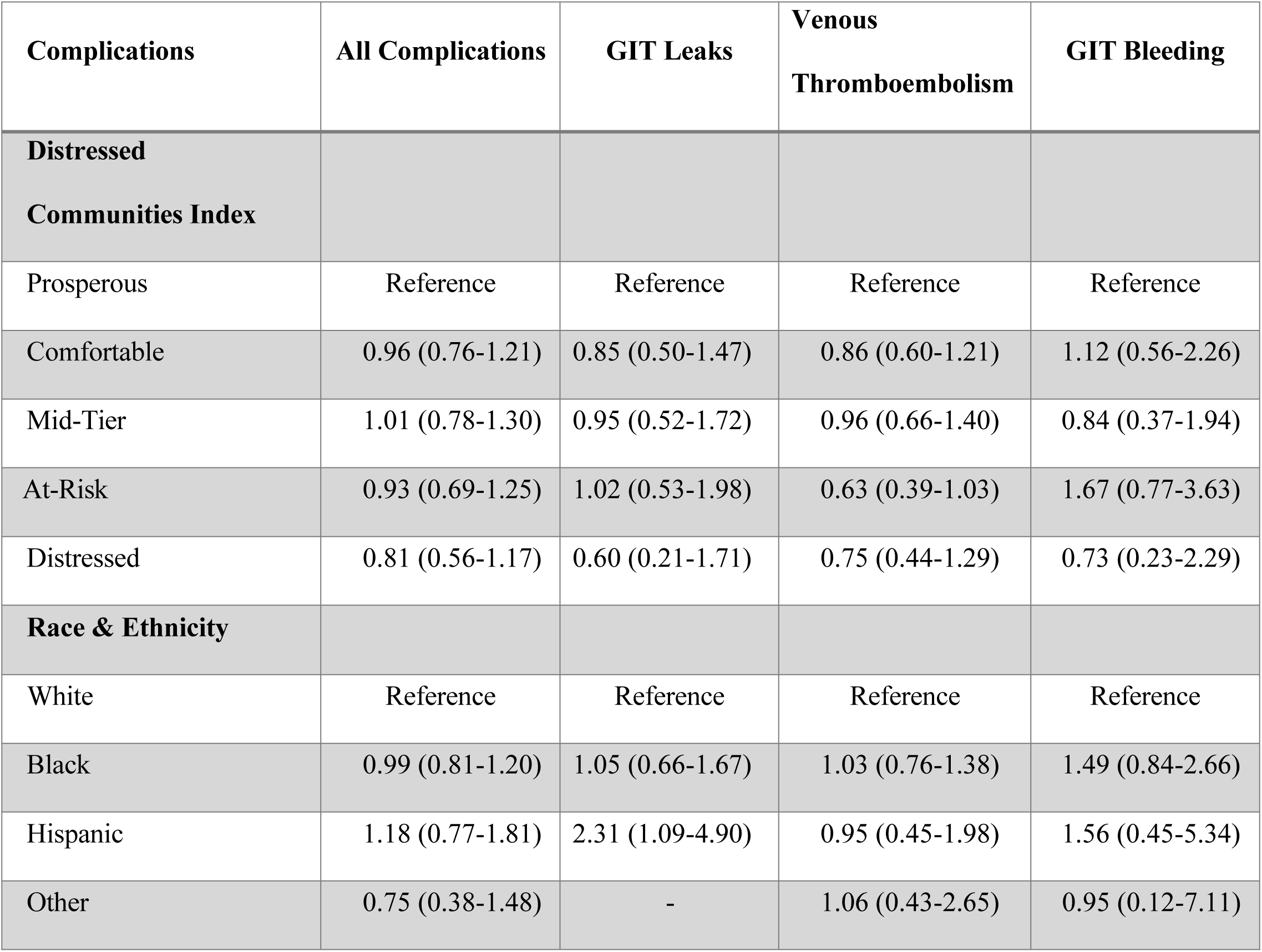
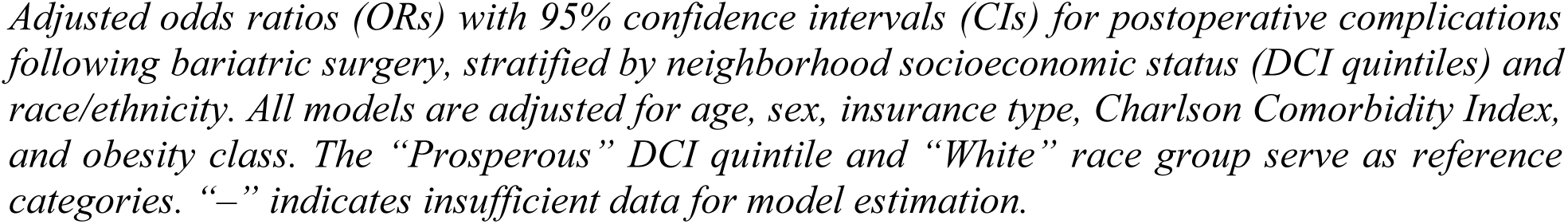
Adjusted Odds of Post-op Complications following Bariatric Surgery by DCI Quintiles and Race & Ethnicity, Maryland 2018–2020.

With regard to specific complications, no DCI quintile demonstrated significantly higher or lower adjusted odds of gastrointestinal (GI) leaks, venous thromboembolism (VTE), or GI bleeding compared to Prosperous communities. Individuals from Distressed areas had lower odds of GI leaks (aOR: 0.60; 95% CI: 0.21–1.71) and GI bleeding (aOR: 0.73; 95% CI: 0.23– 2.29), though these results were not statistically significant. Interestingly, patients from At-Risk communities had a lower odds of VTE (aOR: 0.63; 95% CI: 0.39–1.03), which approached statistical significance (Table 3).

Regarding race and ethnicity, adjusted odds of overall complications did not significantly differ between Black (aOR: 0.99; 95% CI: 0.81–1.20), Hispanic (aOR: 1.18; 95% CI: 0.77–1.81), or Other race (aOR: 0.75; 95% CI: 0.38–1.48) individuals when compared to White patients. However, Hispanic patients had significantly higher adjusted odds of experiencing GI leaks (aOR: 2.31; 95% CI: 1.09–4.90), suggesting a potential disparity in this subgroup. No other statistically significant differences were observed in the rates of specific complications by race or ethnicity (Table 3).

### Race/Ethnicity (Black vs White) and Postoperative Complications across Community Distress Levels

The marginal effects analysis assessed whether race (Black vs. White) moderated the relationship between neighborhood distress and postoperative complications following bariatric surgery. Across all DCI quintiles, the estimated differences in complication rates between Black and White patients were small and statistically non-significant, with confidence intervals crossing zero (Table 4).

**Table 4:**
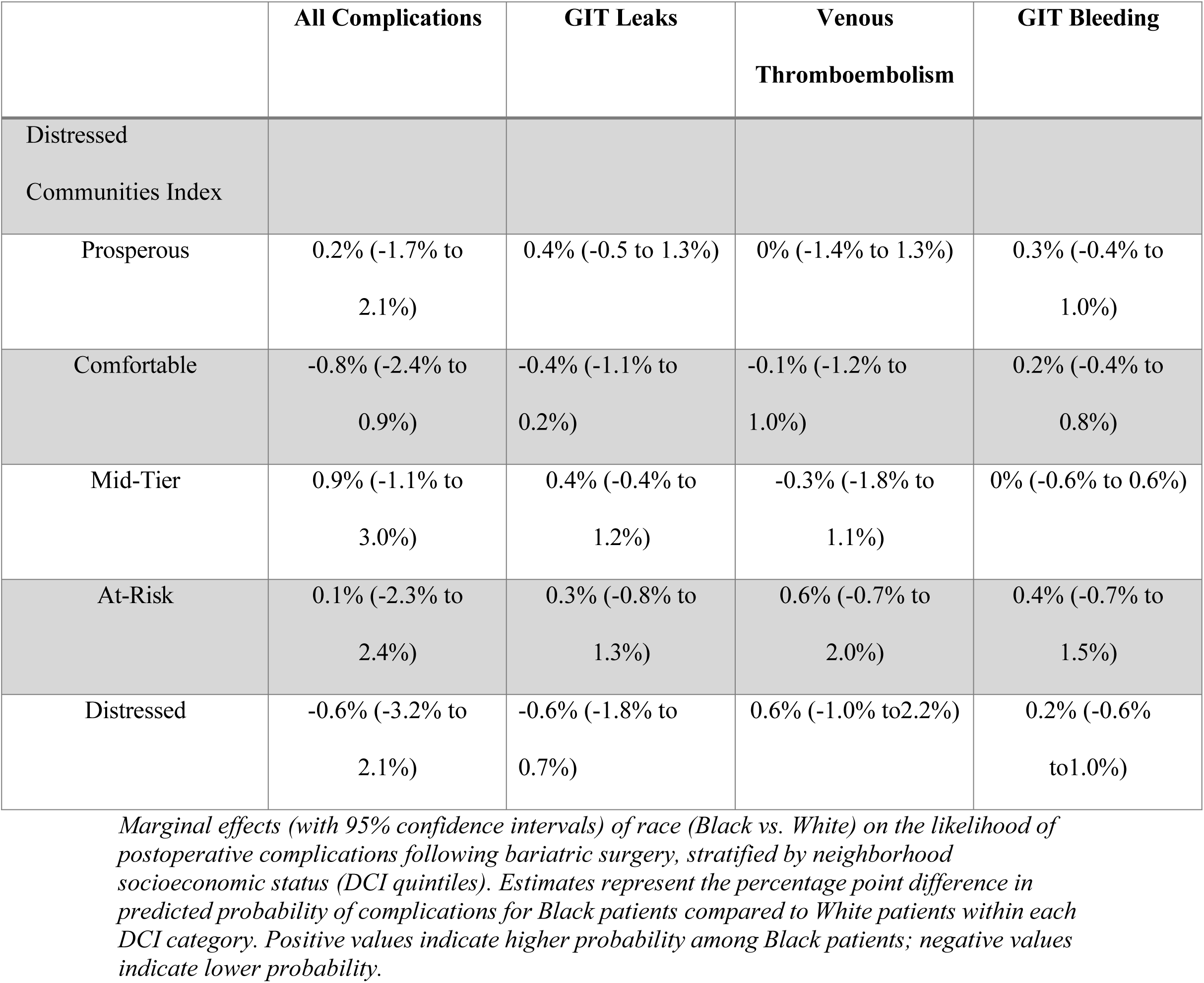
Marginal Effects of Race (Black vs. White) on Post-Op Complications following Bariatric Surgery by DCI, Maryland 2018–2020.

For overall postoperative complications, the marginal differences ranged from −0.8% in Comfortable communities (95% CI: −2.4% to 0.9%) to 0.9% in Mid-Tier communities (95% CI: - 1.1% to 3.0%), indicating no meaningful racial disparities by neighborhood socioeconomic status. Similar patterns were observed for individual complications. For gastrointestinal (GI) leaks, the largest observed difference was 0.4% in Prosperous and Mid-Tier communities, with all estimates falling within non-significant ranges (Table 4).

Venous thromboembolism rates showed a marginal increase of 0.6% for Black patients in both At-Risk and Distressed communities, though these differences were also not statistically significant. Likewise, GI bleeding exhibited negligible differences, with marginal effects close to zero across all quintiles.

Overall, the results suggest that race does not significantly moderate the relationship between community distress levels and short-term complications following bariatric surgery (Table 4).

### Other Race/Ethnicity (Hispanic, Other vs White) and Postoperative Complications Across Community Distress Levels

Marginal effects analysis was conducted to evaluate whether Hispanic and Other race/ethnicity moderated the association between neighborhood distress and composite postoperative complications following bariatric surgery. Across all DCI quintiles, there were no statistically significant differences in complication rates between Hispanic and White patients (Table 5). The marginal effect estimates ranged from −1.6% in Distressed communities (95% CI: −8.4% to 5.2%) to 2.1% in Comfortable communities (95% CI: −2.7% to 6.9%), with all confidence intervals crossing zero (Table 5).

**Table 5:** Marginal Effects of Race (Hispanic, Other vs. White) on Post-Op Complications following Bariatric Surgery by DCI, Maryland 2018–2020.

|  |  | **All Complications** |
| --- | --- | --- |
| **Hispanic vs. White** | **Distressed**<br>**Communities Index** |  |
|  | Prosperous | 1.1% (-4.0% to 6.3%) |
|  | Comfortable | 2.1% (-2.7% to 6.9%) |
|  | Mid-Tier | -0.1% (-4.2% to 4.0%) |
|  | At-Risk | 1.9% (-4.8% to 8.5%) |
|  | Distressed | -1.6% (-8.4% to 5.2%) |
| **Other vs. White** | **Distressed**<br>**Communities Index** |  |
|  | Prosperous | -4.2% (-6.9% to -1.5%) |
|  | Comfortable | -0.4% (-5.4% to 4.6%) |
|  | Mid-Tier | 0.9% (-7.1% to 9.0%) |
|  | At-Risk | -0.6% (-9.1% to 7.9%) |
|  | Distressed | 5.0% (-13.1% to<br>23.1%) |

*Marginal effects (with 95% confidence intervals) of race (Black vs. White) on the likelihood of experiencing any postoperative complication following bariatric surgery, stratified by neighborhood socioeconomic status (DCI quintiles). Estimates represent the percentage point difference in the predicted probability of complications for Black patients compared to White patients within each DCI category. Positive values indicate a higher probability among Black patients, and negative values indicate a lower probability. Marginal effects for individual complications were not reported due to limited sample size.*

For individuals categorized as Other race, results were similarly non-significant in most quintiles, except in Prosperous communities, where Other race patients had significantly lower odds of complications compared to White patients (marginal effect: −4.2%, 95% CI: −6.9% to - 1.5%). In all other DCI quintiles, the marginal effects for other race ranged from −0.6% to 5.0%, with wide confidence intervals that included zero, indicating no meaningful differences. The interaction analyses for Hispanic and Other race groups were limited by small sample sizes, particularly in the distressed quintile, which may have reduced statistical power to detect differences (Table 5).

## DISCUSSION

In this study, we found that only 5.3% of individuals undergoing bariatric surgery had a post-operative complication. We did not observe a significant association between nSES, as measured by the DCI, and the risk of experiencing short-term postoperative complications following bariatric surgery. This finding persisted regardless of whether short-term post-operative complications was analyzed as a composite measure aggregating all short-term postoperative outcomes or as individual complications such as gastrointestinal leaks, venous thromboembolism, and gastrointestinal leaks. We were also unable to identify any moderating effect of race/ethnicity on the association between neighborhood socioeconomic status and short-term postoperative complications, as the relationship remained consistent across racial and ethnic groups, with no significant differences observed.

The distribution of short-term post-operative complications following bariatric surgery revealed that the vast majority of patients (94.7%) experienced no complications, while 5.3% had one or more complications. Among specific complications, gastrointestinal leaks occurred in 0.9% of patients, venous thromboembolism in 2.2%, and gastrointestinal bleeding in 0.6%. In-hospital mortality was rare, occurring in only 0.2% of cases.

These findings highlight that while bariatric surgery is generally associated with low complication rates, certain post-operative risks remain, particularly venous thromboembolism and gastrointestinal leaks, which may warrant closer monitoring [23, 49]. The low incidence of in-hospital mortality aligns with prior research indicating the safety of bariatric procedures in well-selected patients [50, 51]. However, the presence of even a small percentage of severe complications underscores the importance of timely post-operative surveillance and management, particularly among patients with higher baseline risk factors [34, 52].

The lack of statistical significance suggests that the socioeconomic characteristics of the neighborhoods where patients reside may not be a strong determinant of early postoperative complications after bariatric surgery. One potential explanation for this null finding is the relatively low proportion of patients who experienced these complications, which may have resulted in an underpowered analysis. Given that short-term postoperative complications are uncommon following bariatric surgery, our study may have lacked the statistical sensitivity to detect subtle associations. Additionally, it is possible that nSES is not directly related to the immediate post-surgical period in the same way it influences long-term health outcomes. Patients undergoing bariatric surgery often receive standardized perioperative care protocols that may mitigate disparities in short-term complications. Furthermore, hospital-level factors, such as adherence to enhanced recovery after surgery (ERAS) protocols and institutional quality measures, may play a larger role in determining short-term outcomes than neighborhood socioeconomic conditions. These factors could collectively attenuate the expected association between nSES and early post-surgical complications. This finding suggests that the overall safety of bariatric surgery in the short term may, in part, be influenced by standardized perioperative protocols and advancements in surgical techniques. However, further research is needed to determine whether these factors fully explain the low incidence of complications across diverse patient populations.

Beyond statistical power limitations, several mechanisms may contribute to the lack of association between nSES and postoperative complications. Maryland’s unique All-Payer Model, which standardizes hospital reimbursement rates across all patient populations, likely plays a significant role in reducing healthcare disparities [53]. This payment model ensures more equitable access to high-quality bariatric surgery and perioperative care, thereby minimizing variations in outcomes based on neighborhood socioeconomic status [54]. In addition, the existence of high-volume bariatric surgery centers in Maryland, which adhere to standardized perioperative protocols, may have further mitigated disparities in early postoperative complications [55]. Many of these centers implement enhanced recovery after surgery (ERAS) pathways, which include preoperative risk stratification, perioperative infection control measures, and thromboembolism prophylaxis [56]. Such standardization in care delivery likely reduces the influence of socioeconomic factors on immediate postoperative outcomes.

Another plausible explanation for the non-significant associations observed in our study is selection bias. Patients residing in lower socioeconomic status (SES) neighborhoods who successfully undergo bariatric surgery may represent a more resilient and health-conscious subset of their communities. These individuals may have better access to healthcare resources, stronger social support, or higher health literacy compared to others in similarly disadvantaged neighborhoods, potentially attenuating the expected association between neighborhood SES and post-operative complications [17]. These individuals may have greater health literacy, better engagement in preoperative optimization programs, and stronger social support systems, all of which could mitigate the negative effects of neighborhood deprivation on surgical outcomes [17, 57]. This phenomenon, often referred to as the "healthy surgical candidate" effect, suggests that the most disadvantaged patients, who might be at the highest risk for poor outcomes, may not be included in the study due to barriers in accessing surgery in the first place [58]. Consequently, the observed cohort may not fully capture the range of disparities that exist in the broader population.

It is also important to consider whether the DCI and ADI indices, while widely used measures of neighborhood socioeconomic status, may not fully capture the specific social determinants of health such as social support, housing stability, or access to culturally competent care that may have more of a direct influence on postoperative outcomes [59, 60]. For example, individual-level SDOHs such as patient education, access to transportation, health literacy, or adherence to postoperative care plans may better reflect the specific challenges that influence surgical recovery and continuity of care [61]. It is possible that such unmeasured factors play a more critical role in shaping short-term surgical outcomes than the broader neighborhood characteristics captured by the DCI and ADI [62, 63]. DCI and ADI do not capture these challenges because these indices primarily assess structural deprivation based on economic and community-level indicators. Future research should explore whether integrating social vulnerability indices that encompass a broader range of health-related socioeconomic risks may yield more nuanced insights into the relationship between nSES and postoperative outcomes.

### Policy Implications

The findings of this study have important implications for healthcare policy and bariatric surgery research. The absence of disparities in short-term postoperative complications suggests that Maryland’s standardized reimbursement policies and hospital quality improvement initiatives may be effectively ensuring equitable perioperative care. This highlights the potential for health policy interventions to mitigate socioeconomic disparities in surgical outcomes, particularly in states that adopt similar models of healthcare delivery. However, while short-term complications did not vary by nSES, previous research suggests that long-term surgical outcomes, including weight loss maintenance, postoperative readmissions, and nutritional deficiencies, may be more vulnerable to socioeconomic disparities [24]. Future studies should investigate whether neighborhood deprivation influences long-term surgical success, postoperative follow-up adherence, and late complications following bariatric surgery.

### Study Limitations

Several limitations should be considered when interpreting our findings. The reliance on administrative data from the Maryland State Inpatient Database introduces potential misclassification bias, as postoperative complications were identified using ICD-10 codes rather than clinical validation. Additionally, administrative databases lack information on post-discharge factors, such as adherence to follow-up visits, engagement in dietary counseling, and access to postoperative support services, all of which could influence long-term outcomes. Another limitation is the low event rate of postoperative complications, which may have limited the statistical power to detect subtle associations between nSES and outcomes. Future studies could enhance statistical power by pooling data across multiple states or using multi-institutional registries to increase sample sizes. Additionally, the observational study design prevents us from inferring causality, as the relationship between nSES and postoperative outcomes may be confounded by unmeasured hospital- and provider-level factors. Finally, the generalizability of our findings is limited to Maryland, given that the state’s unique healthcare financing model may not be representative of other regions with different reimbursement structures.

To build on these findings, future research should investigate long-term postoperative outcomes by socioeconomic status to determine whether disparities emerge over time. Further studies should also explore the role of patient engagement, social support, and healthcare navigation in postoperative recovery, incorporating qualitative methods and patient-reported outcomes to provide a more comprehensive understanding of the barriers faced by disadvantaged patients. Additionally, research should integrate more granular socioeconomic measures, including employment status, food security, and health literacy, to better capture the complex interplay between social determinants and surgical outcomes. Expanding this analysis to other types of surgical procedures may also provide broader insights into the role of neighborhood factors in shaping health outcomes beyond bariatric surgery.

## Conclusion

In conclusion, this study adds to the growing body of literature examining the relationship between neighborhood socioeconomic status and surgical outcomes. Our findings suggest that in Maryland, where healthcare delivery is highly regulated and standardized, nSES does not significantly impact short-term postoperative complications following bariatric surgery. This underscores the potential protective role of equitable healthcare policies and standardized perioperative protocols in reducing disparities in surgical outcomes. While these results are reassuring, they also raise important questions about long-term disparities, patient-level socioeconomic barriers, and the adequacy of existing indices in capturing healthcare-related social risk factors. Addressing these gaps through future research and targeted policy interventions will be essential to ensure that bariatric surgery remains accessible, effective, and equitable for all patients, regardless of socioeconomic background.

## Data Availability

All relevant data are within the manuscript and its Supporting Information files.

## REFERENCES

[1] Dettoni R, Bahamondes C, Yevenes C, et al. The effect of obesity on chronic diseases in USA: a flexible copula approach. Sci Rep 2023; 13: 1831.

[2] CDC. Adult Obesity Facts, https://www.cdc.gov/obesity/adult-obesity-facts/index.html (accessed 15 March 2025).

[3] Powell-Wiley TM, Poirier P, Burke LE, et al. Obesity and Cardiovascular Disease: A Scientific Statement From the American Heart Association. Circulation; 143. Epub ahead of print 25 May 2021. DOI: 10.1161/CIR.0000000000000973.

[4] Global Data. US businesses and employees face staggering $425.5 billion in economic costs from obesity and overweight in 2023, reveals GlobalData, https://www.globaldata.com/media/healthcare/us-businesses-employees-face-staggering-425-5-billion-economic-costs-obesity-overweight-2023-reveals-globaldata/ (accessed 14 March 2025).

[5] Ram Sohan P, Mahakalkar C, Kshirsagar S, et al. Long-Term Effectiveness and Outcomes of Bariatric Surgery: A Comprehensive Review of Current Evidence and Emerging Trends. Cureus. Epub ahead of print 9 August 2024. DOI: 10.7759/cureus.66500.

[6] Courcoulas AP, Yanovski SZ, Bonds D, et al. Long-term Outcomes of Bariatric Surgery: A National Institutes of Health Symposium. JAMA Surg 2014; 149: 1323.

[7] Lee W, Almalki O. Recent advancements in bariatric/metabolic surgery. Ann Gastroenterol Surg 2017; 1: 171–179.

[8] Alsuhibani A, Thompson JR, Wigle PR, et al. Metabolic and Bariatric Surgery Utilization Trends in the United States: Evidence From 2012 to 2021 National Electronic Medical Records Network. Ann Surg Open 2023; 4: e317.

[9] Alalwan AA, Friedman J, Park H, et al. US national trends in bariatric surgery: A decade of study. Surgery 2021; 170: 13–17.

[10] Davey MG, Donlon NE, Fearon NM, et al. Evaluating the Impact of Enhanced Recovery After Surgery Protocols on Surgical Outcomes Following Bariatric Surgery—A Systematic Review and Meta-analysis of Randomised Clinical Trials. Obes Surg 2024; 34: 778–789.

[11] Gulinac M, Miteva DG, Peshevska-Sekulovska M, et al. Long-term effectiveness, outcomes and complications of bariatric surgery. World J Clin Cases 2023; 11: 4504–4512.

[12] Lim R, Beekley A, Johnson DC, et al. Early and late complications of bariatric operation. Trauma Surg Acute Care Open 2018; 3: e000219.

[13] Dorman RB, Miller CJ, Leslie DB, et al. Risk for Hospital Readmission following Bariatric Surgery. PLoS ONE 2012; 7: e32506.

[14] Daigle CR, Brethauer SA, Tu C, et al. Which postoperative complications matter most after bariatric surgery? Prioritizing quality improvement efforts to improve national outcomes. Surg Obes Relat Dis 2018; 14: 652–657.

[15] Haidar S, Vazquez R, Medic G. Impact of surgical complications on hospital costs and revenues: retrospective database study of Medicare claims. J Comp Eff Res 2023; 12: e230080.

[16] Masanam MK, Grossman DA, Neary J, et al. Disparities in the impact of access to and outcomes of bariatric surgery among different ethnoracial and socioeconomic populations: a narrative review of the literature. Ann Laparosc Endosc Surg 2023; 8: 34–34.

[17] Funk LM, Alagoz E, Murtha JA, et al. Socioeconomic disparities and bariatric surgery outcomes: A qualitative analysis. Am J Surg 2023; 225: 609–614.

[18] Miras AD, Le Roux CW. Can medical therapy mimic the clinical efficacy or physiological effects of bariatric surgery? Int J Obes 2014; 38: 325–333.

[19] Chang S-H., Freeman NLB, Lee JA, et al. Early major complications after bariatric surgery in the USA, 2003–2014: a systematic review and meta-analysis. Obes Rev 2018; 19: 529–537.

[20] Mizera M, Wysocki M, Walędziak M, et al. The impact of severe postoperative complications on outcomes of bariatric surgery—multicenter case-matched study. Surg Obes Relat Dis 2022; 18: 53–60.

[21] Marshall JS. Roux-en-Y Gastric Bypass Leak Complications. Arch Surg 2003; 138: 520.

[22] Silva AFD, Mendes KDS, Ribeiro VDS, et al. Risk factors for the development of surgical site infection in bariatric surgery: an integrative review of literature. Rev Lat Am Enfermagem 2023; 31: e3798.

[23] Froehling DA, Daniels PR, Mauck KF, et al. Incidence of Venous Thromboembolism After Bariatric Surgery: A Population-Based Cohort Study. Obes Surg 2013; 23: 1874–1879.

[24] Stenberg E, Näslund I, Persson C, et al. The association between socioeconomic factors and weight loss 5 years after gastric bypass surgery. Int J Obes 2020; 44: 2279–2290.

[25] Liu N, Venkatesh M, Hanlon BM, et al. Association Between Medicaid Status, Social Determinants of Health, and Bariatric Surgery Outcomes. Ann Surg Open 2021; 2: e028.

[26] Fox M. American College of Surgeons. Social determinants of health and surgery: An overview, https://www.facs.org/for-medical-professionals/news-publications/news-and-articles/bulletin/2021/05/social-determinants-of-health-and-surgery-an-overview/ (accessed 15 March 2025).

[27] Holbert SE, Andersen K, Stone D, et al. Social Determinants of Health Influence Early Outcomes Following Lumbar Spine Surgery. Ochsner J 2022; 22: 299–306.

[28] Patrick WL, Bojko M, Han JJ, et al. Neighborhood socioeconomic status is associated with differences in operative management and long-term survival after coronary artery bypass grafting. J Thorac Cardiovasc Surg 2022; 164: 92–102.e8.

[29] Sengupta A, Bucholz EM, Gauvreau K, et al. Impact of Neighborhood Socioeconomic Status on Outcomes Following First-Stage Palliation of Single Ventricle Heart Disease. J Am Heart Assoc 2023; 12: e026764.

[30] Bhaktaram A, Kress AM, Li Z, et al. Unpacking Neighborhood Socioeconomic Status in Children’s Health Research from an Environmental Justice Perspective: A Scoping Review. Curr Environ Health Rep 2024; 11: 288–299.

[31] Pampel FC, Krueger PM, Denney JT. Socioeconomic Disparities in Health Behaviors. Annu Rev Sociol 2010; 36: 349–370.

[32] Levy BL, Vachuska K, Subramanian SV, et al. Neighborhood socioeconomic inequality based on everyday mobility predicts COVID-19 infection in San Francisco, Seattle, and Wisconsin. Sci Adv 2022; 8: eabl3825.

[33] Lusk JB, Blass B, Mahoney H, et al. Neighborhood Socioeconomic Disadvantage and 30-Day Outcomes for Common Cardiovascular Conditions. J Am Heart Assoc 2024; 13: e036265.

[34] Husain F, Jeong IH, Spight D, et al. Risk factors for early postoperative complications after bariatric surgery. Ann Surg Treat Res 2018; 95: 100.

[35] Sheka AC, Kizy S, Wirth K, et al. Racial disparities in perioperative outcomes after bariatric surgery. Surg Obes Relat Dis 2019; 15: 786–793.

[36] Welsh LK, Luhrs AR, Davalos G, et al. Racial Disparities in Bariatric Surgery Complications and Mortality Using the MBSAQIP Data Registry. Obes Surg 2020; 30: 3099– 3110.

[37] Khattab MA, Mohammed ATA, Alqahtani AZM, et al. The Role of Ethnic Disparities in the Outcomes of Bariatric Surgery: A Systematic Review and Meta-Analysis. Cureus. Epub ahead of print 5 May 2022. DOI: 10.7759/cureus.24743.

[38] Premkumar A, Samaan JS, Samakar K. Factors Associated With Bariatric Surgery Referral Patterns: A Systematic Review. J Surg Res 2022; 276: 54–75.

[39] Agency for Healthcare Research and Quality. Healthcare Cost and Utilization Project (HCUP) User Support (HCUP-US), https://www.ahrq.gov/cpi/about/otherwebsites/hcupnet.ahrq.gov/index.html (accessed 15 March 2025).

[40] Jacobsen HJ, Nergard BJ, Leifsson BG, et al. Management of suspected anastomotic leak after bariatric laparoscopic Roux-en-y gastric bypass. Br J Surg 2014; 101: 417–423.

[41] Dick A, Byrne TK, Baker M, et al. Gastrointestinal bleeding after gastric bypass surgery: nuisance or catastrophe? Surg Obes Relat Dis 2010; 6: 643–647.

[42] Carvalho L, Almeida RF, Nora M, et al. Thromboembolic Complications After Bariatric Surgery: Is the High Risk Real? Cureus. Epub ahead of print 6 January 2023. DOI: 10.7759/cureus.33444.

[43] Ashrafi D, Osland E, Memon MA. Bariatric surgery and gastroesophageal reflux disease. Ann Transl Med 2020; 8: S11–S11.

[44] Glasheen WP, Cordier T, Gumpina R, et al. Charlson Comorbidity Index: ICD-9 Update and ICD-10 Translation. Am Health Drug Benefits 2019; 12: 188–197.

[45] Charlson ME, Carrozzino D, Guidi J, et al. Charlson Comorbidity Index: A Critical Review of Clinimetric Properties. Psychother Psychosom 2022; 91: 8–35.

[46] Comoglu S. Does the Charlson comorbidity index help predict the risk of death in COVID-19 patients? North Clin Istanb. Epub ahead of print 2022. DOI: 10.14744/nci.2022.33349.

[47] Department of Pulmonary Diseases, Faculty of Medicine, Kocaeli University, Kocaeli, Turkey, Argun Baris S, Boyaci H, et al. Charlson Comorbidity Index in Predicting Poor Clinical Outcomes and Mortality in Patients with COVID-19. Turk Thorac J 2022; 23: 145–153.

[48] Sjöholm K, Anveden Å, Peltonen M, et al. Evaluation of Current Eligibility Criteria for Bariatric Surgery. Diabetes Care 2013; 36: 1335–1340.

[49] Lim R, Beekley A, Johnson DC, et al. Early and late complications of bariatric operation. Trauma Surg Acute Care Open 2018; 3: e000219.

[50] Moulla Y, Lyros O, Blüher M, et al. Feasibility and Safety of Bariatric Surgery in High-Risk Patients: A Single-Center Experience. J Obes 2018; 2018: 1–6.

[51] Pories WJ. Bariatric Surgery: Risks and Rewards. J Clin Endocrinol Metab 2008; 93: s89–s96.

[52] Neuberg M, Blanchet M-C, Gignoux B, et al. Connected Surveillance for Detection of Complications After Early Discharge from Bariatric Surgery. Obes Surg 2020; 30: 4669–4674.

[53] Maryland Hospital Association. The Maryland Model, https://mhaonline.org/caring-for-communities/the-maryland-model/ (accessed 15 March 2025).

[54] Frederick Health. Enhanced Recovery After Surgery Program (ERAS), https://www.frederickhealth.org/services/surgical-care/enhanced-recovery-after-surgery-program-eras-/ (accessed 15 March 2025).

[55] Awad S, Carter S, Purkayastha S, et al. Enhanced Recovery After Bariatric Surgery (ERABS): Clinical Outcomes from a Tertiary Referral Bariatric Centre. Obes Surg 2014; 24: 753–758.

[56] Huh Y-J, Kim DJ. Enhanced Recovery after Surgery in Bariatric Surgery. J Metab Bariatr Surg 2021; 10: 47.

[57] Mahoney ST, Strassle PD, Farrell TM, et al. Does Lower Level of Education and Health Literacy Affect Successful Outcomes in Bariatric Surgery? J Laparoendosc Adv Surg Tech 2019; 29: 1011–1015.

[58] Ziegelmann M, Köhler TS, Bailey GC, et al. Surgical patient selection and counseling. Transl Androl Urol 2017; 6: 609–619.

[59] Vassilaki M, Petersen RC, Vemuri P. Area Deprivation Index as a Surrogate of Resilience in Aging and Dementia. Front Psychol 2022; 13: 930415.

[60] Ghirimoldi FM, Schmidt S, Simon RC, et al. Association of Socioeconomic Area Deprivation Index with Hospital Readmissions After Colon and Rectal Surgery. J Gastrointest Surg 2021; 25: 795–808.

[61] De Oliveira GS, McCarthy RJ, Wolf MS, et al. The impact of health literacy in the care of surgical patients: a qualitative systematic review. BMC Surg 2015; 15: 86.

[62] Theiss LM, Wood T, McLeod MC, et al. The association of health literacy and postoperative complications after colorectal surgery: A cohort study. Am J Surg 2022; 223: 1047–1052.

[63] Stephens CQ, Yap A, Vu L, et al. Comparative Analysis of Indices for Social Determinants of Health in Pediatric Surgical Populations. JAMA Netw Open 2024; 7: e2449672.

